# A new Reproduction Index *R*_*i*_ and its Usefulness for Germany’s Covid19-Data

**DOI:** 10.1101/2021.03.29.21254581

**Authors:** Robert N.J. Conradt

## Abstract

In the course of a large-scale infectious disease a time-dependent Reproduction rate is an important parameter for political, economic and social decisions. In this paper we focus on that parameter and introduce a mathematical implementation in addition to the mostly used definition of Robert-Koch-Institute (RKI) in Germany.

Such value is of particular interest in order to serve as a criterion for possible Lock-Downs and “LockUps” in society and can provide deep insights into a pandemic event.

Both the definition of the new Reproduction index and the RKI’s Reproduction number are compared analytically, applied to simple model calculations and finally on real Covid19 data. Clear advantages of the new Reproduction index become apparent and some weaknesses of the RKI’s Reproduction number become clearly visible.

In addition we propose two additional ways of displaying pandemic data to have the pandemic behaviour at a glance. We find that some signatures of the pandemic appear now very well expressed - especially in conjunction with the new Reproduction index R_i_.

This all could be very helpful for future political, social and economic decisions.

## 1 Introduction

The Reproduction index indicates how many people are infected by another infected person during their infectious phase, if such infected person appears in the population of susceptible individuals with a given social environment. The Reproduction index is a dimensionless number. For a population of infected individuals with *R*_*i*_ *>* 1 this means that the part of infected individuals increases. For *R*_*i*_ = 1 the size of this infected sub-population stays constant and for *R*_*i*_ *<* 1 it shrinks - the pandemic is fading out.

All these properties are also claimed by the definition of the well known RKI-Reproduction number *R*_*t*_ [8].^2^

In Fig.1, the necessary states for an infection and the transition rates between the single states can be represented using a simple SIRD-model.

**Figure 1:**
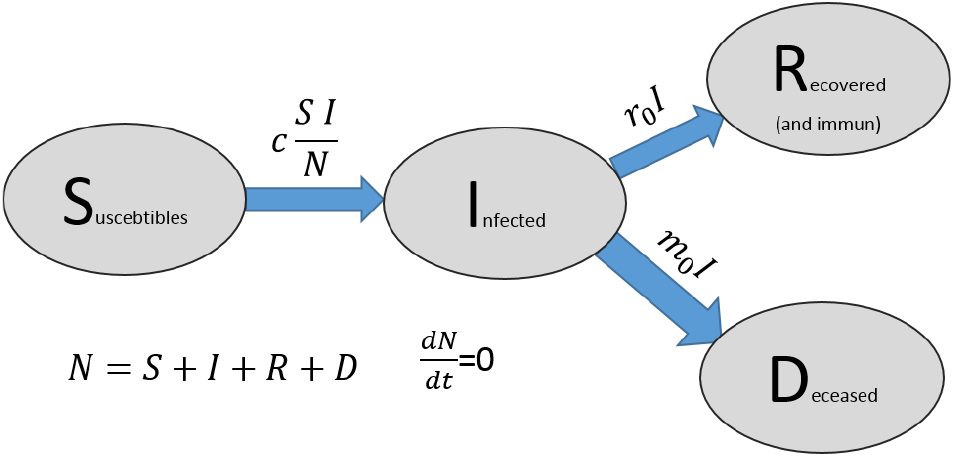
Simple SIRD-model: Between the states S (Susceptible), I (Infected), R (Recovered) and D (Deceased), individuals change their state using the specified rates. N is the total number of all individuals and remains constant.

The transition rates 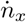 from one state of the model to another are given by the following set of expressions:

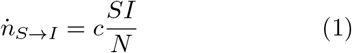

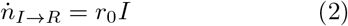

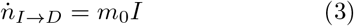

Applied to the model given in Fig.1 this results in the well-known set of differential equations:

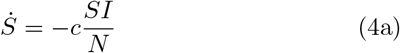

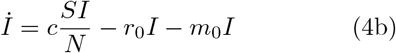

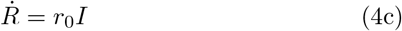

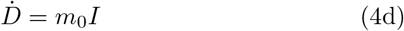

*Remarks:* In Eq.(1), the tools to contain a pandemic are easily identified as factors of the product *c · I · S*: Reducing *c* can be achieved with measures such as masks, hygiene, contact reduction. *S* is reduced actively by vaccination^3^ and *I* can be reduced by identifying infected persons (testing, tracing). Since all measures come in as a product, they are equally important - but vary in ease of use.

The much more detailed compartment model given by Meyer-Hermann’s group [5] knows significantly more states and is important for a detailed understanding of the whole infection process. It can primarily distinguish between identified infected and unrecognized infected individuals. To bring this model to full value it is required to calibrate the number of undetected (asymptomatic) infected people against the number of obviously infected people. As long as there are no large-scale tests on representative numbers of the population this remains an issue. Unfortunately, currently this is not done systematically in Germany.

Streeck et al. did this on a small scale in the well known “Heinsberg-study” [11]. The high value of this study consists of the response to a well defined pulse-like superspreading-event.

Using Fig.1 and Eq.(4) the definition of a Reproduction index *R*_*i*_ becomes immediately clear and simple. It results from the ratio of new infections (incidence) to the average number of active infections (prevalence) for the duration of infectivity Δ*t*:

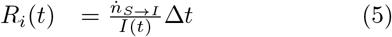

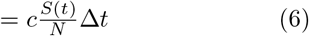

Δ*t* is the dwell time in the Infected state and can also be represented by (*r*_0_ + *m*_0_)^*−*1^. Eq.(6) thus becomes

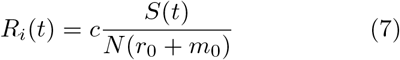

In words: the number of new infections *R*_*i*_(*t*) caused by an infected person depends on the (time dependant) probability to meet susceptible persons and on the “efficiency” of the contact (from the point of view of the virus)^4^ between the infected person and susceptible person. The duration of the infectiosity (*r*_0_ + *m*_0_)^*−*1^ ofcourse plays a role. Notice: the number of new infections caused by an infected person does not depend on how many other infected persons exist around the infecting person.

Using *S*(*t* = 0) = *N*, the basic Reproduction index *R*_*i*,0_ is obtained from Eq.(7):

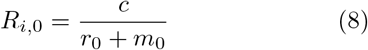

Using Eq.(7) in Eq.(4b) we immediately obtain

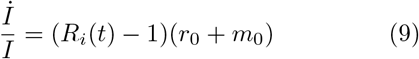

The pandemic stagnates at a critical reproduction index *R*_*i,c*_ = 1 corresponding to *İ* = 0. Above this, the number of infected individuals increases and for values *R*_*i*_(*t*) *<* 0 the pandemic fades away.

For the period and duration Δ*t* of infectivity, we draw on a distribution given by Cori et al. [3] for the SARS 2003 pathogen. This agrees well with the analysis of a smaller sample of the first cases from Wuhan, China, by Li et al. [6].

For the duration Δ_*t*,90%_ of infectiosity resulting from Fig.2 we obtain about 12.3 days.

**Figure 2:**
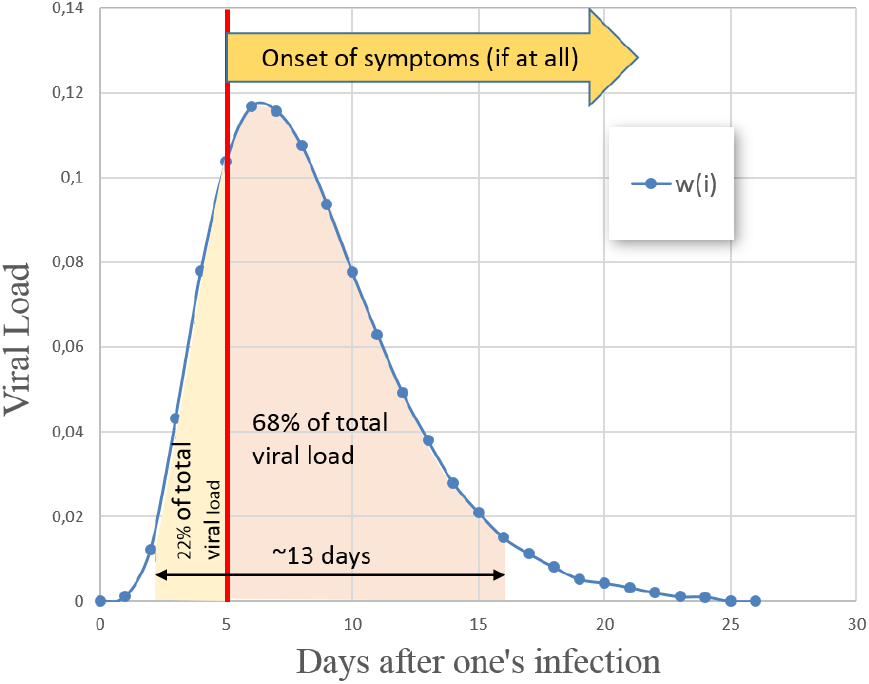
Distribution of infectivity w_i_ in SARS-2003. After an infection at t = 0, the infectivity, i.e., the time course of the discharge of viruses, is as shown by Cori et. al. [3]. The colored area shows the 90%-quantil of infectiosity. In an interval Δt_90_ = [3, 2…15, 5], 90% of the total virus-load emitted by an infected person is released. Please note that 22% of the virus-load is spread before any symptoms show.

This coincides quite well with the inverse of the rate *r*_0_ + *m*_0_ which is 14.2 days and corresponds to the dwell time in the “Infected” state.

To conclude the introduction, we present a numerical integration of the system of equations (4) together with real data of the same time.

Apart from the simple accessibility in a simulation, values for 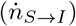 and (*I*) are given in the Daily Situation Reports of the Robert-Koch-Institute (RKI)[9] ^5^.

The following parameters belonging to Fig.3 describe the first wave 2020 very well^6^:

**Figure 3:**
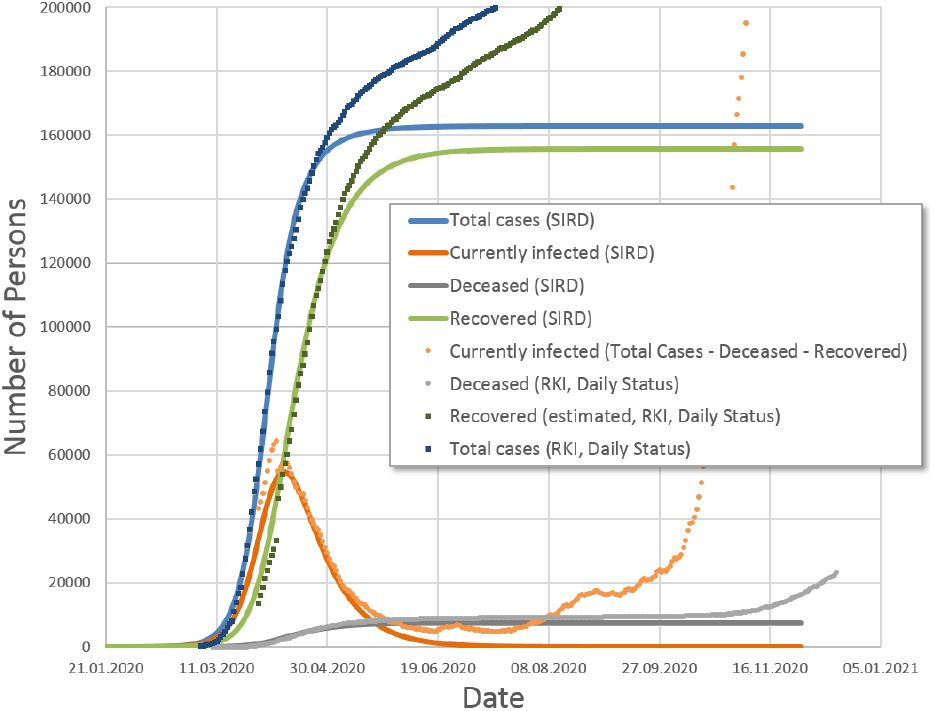
Course Covid19. Data of the pandemic from the RKI Daily Situation Reports [9]. The solid curves (simulation according to Eq.(4)) decsribe very well the course of the first wave. The continuation of the pandemic is already indicated at the end of April when the true numbers leave the simulation-curves.

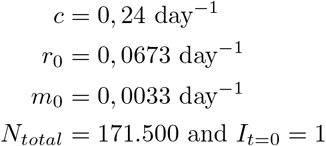

## 2 Definitions of a Reproduction Rate

Let us look at the properties of the new Reproduction index *R*_*i*_ as given in Eq.(5) and the Reproduction number *R*_*t*_ given by the RKI in terms of properties and differences.

### 2.1 Proposal of a supplementary Reproduction Index *R*_*i*_

Instead of using the continuous version according to Eq.(5) we have to use the daily given numbers^7^ by RKI [9] and define a discrete version of Eq.(5). In Eq.(10), we average the values of incidence over 3 days with the actual day in the middle to get a little smoothing and divide by the mean of the prevalences before. The mean value of the prevalence could be obtained by averaging the number of active cases for about 10 days, which in turn is in the same order of magnitude as the duration of the infectivity. Since the variation in prevalence is not as strong as in incidence-values, finally the choice of the interval for averaging prevalence plays a minor role.

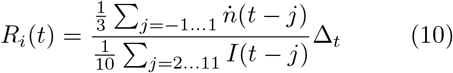

For Δ_*t*_, we used an approximate value of 13 days for the duration of infectivity in the evaluations given below^8^.

### 2.2 *R*_*t*_ given by the Robert-Koch-Institute

The RKI reproduction number *R*_*t*_ is based on the idea of calculating the ratio of the incidence values of two successive instants. Such a snapshot can itself consist of an average over a few days for smoothing. The time distance between these two snapshots is the generation time *t*_*G*_. This is the time interval between one’s own infection and a “successful” transmission of the infection to a third person. The RKI uses a value of 4 days for *t*_*G*_. First we define a continuous version for *R*_*t*_ to check its properties applied on the DEs Eq.(4). As term for incidence we use 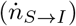 as given in Fig.1 and Eq.(1).

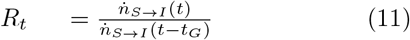

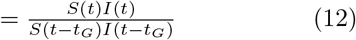

Please note that this has no correspondence to Eq.(5) or Eq.(6). First of all it is surprising that *R*_*t*_ does not depend on *c*, although this is generally considered to be the parameter for the coupling between susceptibles and infected. It is the parameter that is also supposed to be reduced by social-distancing as a measure. Thus, such a measure would not be able to be seen in RKI’s *R*_*t*_.

In a first-order-approximation, we get 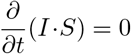 as a condition for the critical Reproduction number *R*_*t,c*_ = 1. This differs from the correspondig condition *İ* = 0 for *R*_*i,c*_ = 1 and results in a slightly different timing when crossing then *R* = 1-line.

Also Eq.(12) can be written in a discrete version with daily case-numbers. The ratio of the mean-value of new cases of two subsequent time intervals is determined. RKI uses an averaging over 4 days for each of the two intervals and a shift of *t*_*G*_ =4 days. In the following graph Fig.4 the slope of the total number of cases corresponds to the (averaged) number of new cases. The ratio between two subsequent slopes is RKI’s *R*_*t*_.

**Figure 4:**
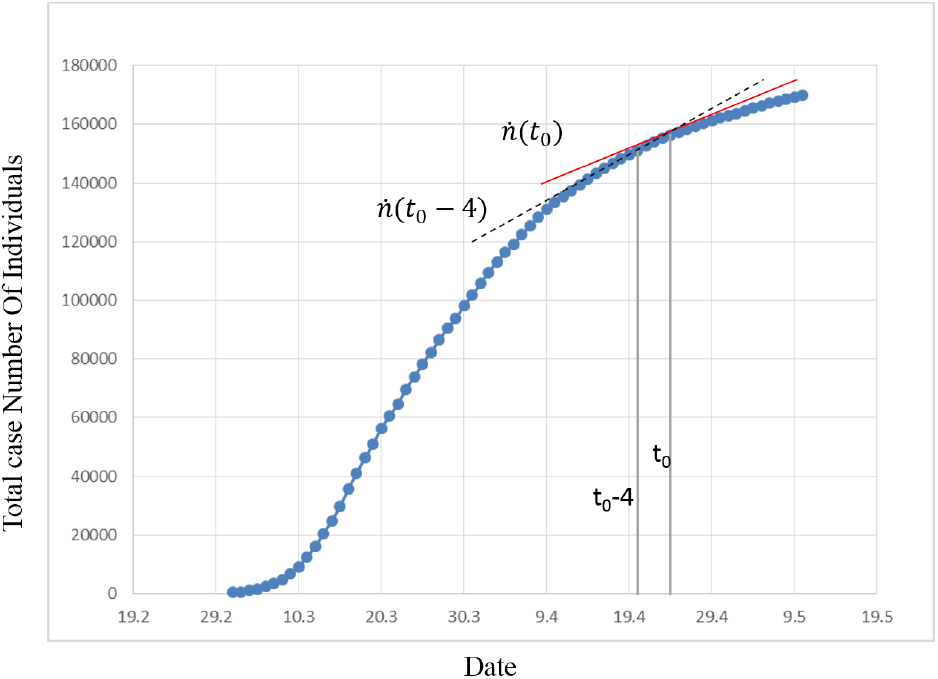
For the definition of R_t_ by the RKI: Shown are the cumulative case numbers announced daily by the RKI [8]. The Reproduction number R_t_ is given as the ratio of the two averaged slopes plotted.

Assuming that the time axis is in discrete steps (days), the following definition is given for the Reproduction number *R*_*t*_ [7][1]:

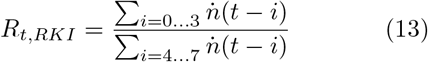

With Eq.(13), the relative change in the averaged case counts of two consecutive intervals of 4 days each is given^9^.

Let us summarize some very remarkable properties of the definitions according to Eq.(12)or Eq.(13):

- *R*_*t*_ does not depend on contact-intensity. The reproduction number is independent of whether infectious persons are in the vicinity of suscebtible persons or not. According to this, quarantine measures would have no effect in *R*_*t*_ and thus can not be monitored by that definition of *R*_*t*_.
- As example, 10 new infections in each of 8 consecutive days yield a value *R*_*t*_ = 1, just as 10.000 new infections in each of 8 consecutive days result in a value of *R*_*t*_ = 1, too; regardless by how many currently infected persons these new infections were generated. 10 new infections generated by (e.g.) 1000 active cases would be acceptable while 10.000 new infections generated by 1000 infected would be very critical.
- The definition according to Eq.(13) reflects the relative change of the new infection rate, it does not represent a Reproduction rate in the sense of Eq.(5) or Eq.(6) in section 1 as the number of infected is ignored.
- Even when the infection has almost completely faded away, the value *R*_*t*_ stays close to “1”.
- Since only case-numbers are required, the calculation of *R*_*t*_ is is done quite easy.

### 2.3 Properties of the two Reproduction Rates

To compare the implementations of the two methods of calculating *R*_*t*_ and *R*_*i*_, the definitions according to Eq.(13) and Eq.(10) are applied on the simulation-dataset shown in Fig.3 (solid lines). Here we can test the two definitions of a Reproduction rate on a model system before applying them in a second step to real Covid19 data (dotted curves in Fig.3) to gain new insights there.

#### 2.3.1 *R*_*t*_ and *R*_*i*_ in Simulation-Data

Let us first have a look at the definitions of *R*_*t*_ and *R*_*i*_ and their application to the simulation-dataset already shown in Fig.3:

It can be clearly seen in Fig.5 that the course of a pandemic wave is reproduced completely differently by the two reproduction rates.

**Figure 5:**
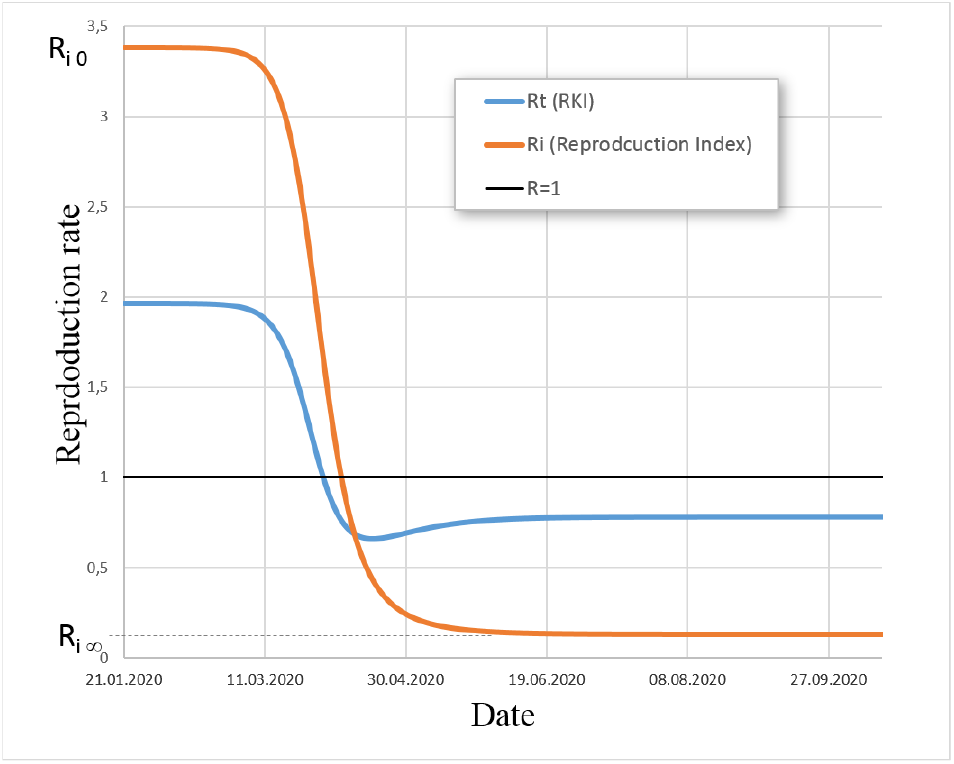
R_t_ associated with SIRD simulation. The comparison of the two calculation methods for R_t_ in contrast. We notice the qualitative difference that does not show the RKI R-curve to decay monotonically towards the end of the pandemic. The time at which R_i_ and R_t_ indicate a stationary situation (ReproductionRate = 1) differs slightly.

In contrast to *R*_*t*_, the alternative calculation *R*_*i*_ starts at *R*_0_ and falls monotonically. Exactly when *İ* = 0 the index *R*_*i*_ takes the value “1” on 9.4.2020^10^. Here New cases and (Recovered + Deceased) balance each other. Then *R*_*i*_ tends towards a value *R*_*∞*_, which is given in Fig.14 (Supporting Information). For *R*_0_ = 3, 3 we find *R*_*i,∞*_ = 0, 13.

#### 2.3.2 *R*_*t*_ and *R*_*i*_ applied on real Data

Applying the definitions for *R*_*t*_ acc. Eq.(13) and for *R*_*i*_ given by Eq.(10) on real Covid19-data is shown in Fig.6^11^.

**Figure 6:**
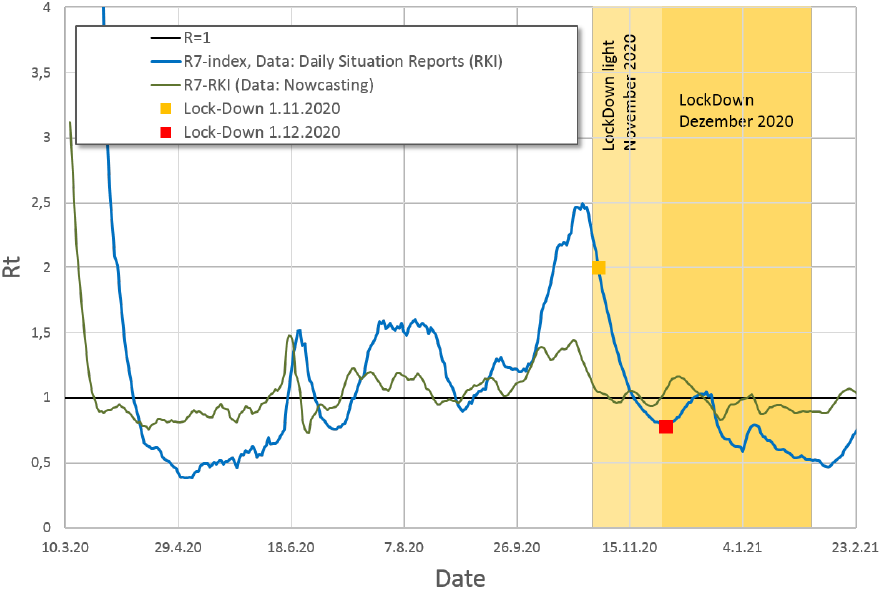
Comparison of Reproduction rates. Interesting is the property of R_t,RKI_ to tend to the value “1”, while R_i_ is able to show values over a significantly wider range.

Clearly a difference can be seen when applying the two definitions of the Reproduction rate to the real data.

Easily seen is the tendency of RKI’s definition of *R*_*t*_ to gravitate toward the value “1”.

In contrast the definition of *R*_*i*_ proposed here has the property of reflecting the dynamic event more pronouncedly. The intermediate increase of *R*_*i*_ up to a value of 2.5 is not shown by RKI’s value *R*_*t*_.

Since the daily values of *R*_*t*_ published by the RKI are based on the data set of the nowcasting [8], for the sake of completeness the small differences to the evaluation based on the daily situation reports are shown in Supporting Information (3.2.4, p.10).

A detailed analysis or interpretation of signatures in Fig.6 is not carried out here, but a few striking signatures should nevertheless be mentioned: the clearly visible outbreak at the Tönnies/Gütersloh company around 18.6.2020, a comparatively high number “imported infections” during 4 weeks startig at the beginning of August and the strong increase of new infections at the beginning of October rising up to a *R*_*i*_ = 2, 5.

At the end of October, a LockDown-Light was imposed in Germany at *R*_*i*_ = 2 while *R*_*i*_ was already decreasing. Since a sharp increase in new infections was expected at this time of the year, the decreased valued *R*_*i*_ = 0.77 at the end of November is remarkable^12^.

Since the case numbers were comparatively high at about 20,000 per day and the Reproduction number of the RKI did not signal any change, a tightening of the lock-down was announced for December 2020 - although *R*_*i*_ was already quite low. This also happened due to the 7th Ad-Hoc-statement of the Leopoldina published on December 8th, 2020 [4]. In particular, this statement formulates a target value for *R* = 0.7 … 0.8.

By such criteria, any Reproduction rate is given an importance that it should necessarily be able to satisfy.

### 2.4 Proposal for two new kind of Plots

It is easy to consider that two projections of the three-dimensional SI(R+D) space fully describe the pandemic. Here we present two projections of that three-dimensional space with slightly different variables^13^. We have choosen the coordinate-system (*R*_*i*_ | Incidence | Prevalence) for the following two plots because they are widely spread and give a quite good insight respectively overview over the pandemic - and they fully describe the pandemic. Please note: the plots don’t explain the pandemic in the meaning that one can fully understand the pandemic - they describe some details and give the the required information for decisions at a glance. We think that the projection of the 3-dimensional space only on the axis “Incidence” is not sufficient since important information stays invisible.

#### 2.4.1 Incidence vs. Prevalence

In the first type of plot we focus on the state of the pandemic by using “Incidence vs. Prevalence” as coordinate-system which can be regarded as something like the state diagram of a pandemic:

In Fig.7, the most important curve is the orange dotted-lined trajectory^14^ of new infections (7-days averaged) vs. active/infected cases at the same time.

**Figure 7:**
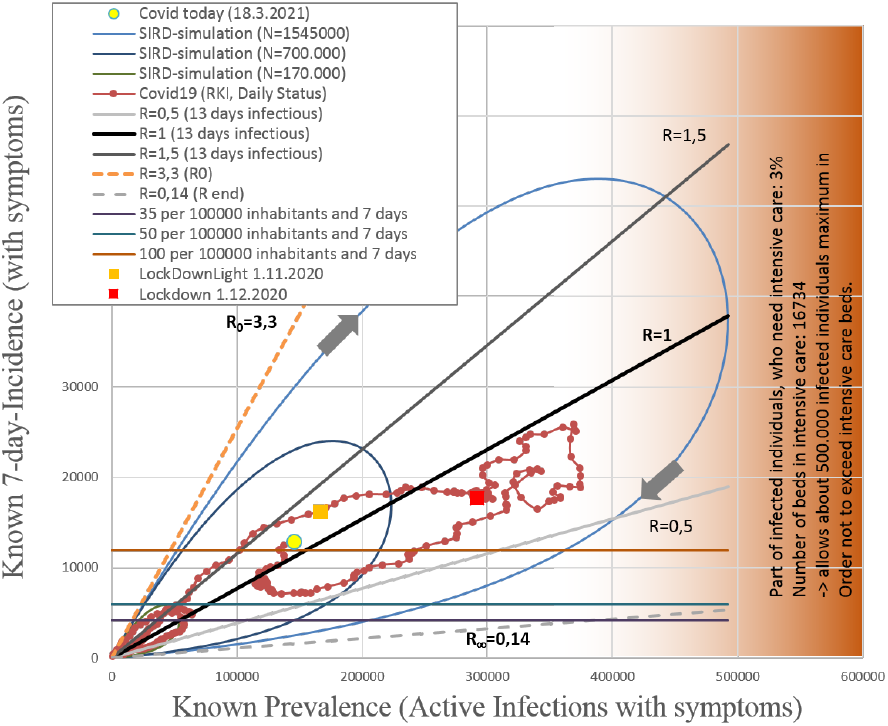
The “Pandemic state Compass”. provides a state diagram of a pandemic event by plotting incidences vs. prevalences. The orange dot-line-curve (Covid19 (RKI, daily Status)) shows true data. Additional guidelines are given for different Reproduction indices (lines through origin), 7-day-incidences (horizontal lines, used currently for political decisions), health-care limit region (vertically oriented region at the right side) and free running pandemic waves (blobs for different numbers of involved individuals).

We can recognize the first wave 2020 as nearly free running pandemic event (blob-shaped at bottom left corner), while the second wave with its numbers has almost reached the capacity of the German health care system.

Further entry details observable in Fig.7:

- Lines through origin represent different Reproduction indices, whose slope is given by Eq.(5)^15^.
- The 7-day incidences which serve currently in politics as criteria can be marked as horizontal lines^16^.
- Based on data from Stang et al. [10] there are 16734 intensive care beds available in Germany ^17^. Furthermore, the percentage of patients requiring an intensive care bed is 3%. This implies that the maximum number of active cases should not exceed 500,000 - otherwise the health care system reaches its limits. This is indicated by the reddish part in the right side of Fig.7. The closer we come to this region the more the German health care system is stressed. Arentz and Wild [2] give a valuable overview on 15 European states and the capacity of their health-care system.
- As a final guide, blob-shaped simulations of unaffected pandemic waves are drawn. The simulations were calculated according to Eq.(4) with the parameters of the first 2020 wave (see Fig.3). Between the “blobs” only the envolved population size *N*_*ges*_ was varied. This results in additional useful blob-shaped auxiliary lines.

#### 2.4.2 Reproduction Index vs. Incidence

The second very informative visualization is to plot *R*_*i*_ against the incidence. This plot is based on the use of Eq.(10) for *R*_*i*_. It is another projection of the “3-dimensional pandemic space” where the “incidence” axis coincides with that in Fig.7.

We remark that, despite a decreasing incidence, the pandemic wave gains very strong momentum first in spring 2020 after the first wave (where it was strongly depressed by the following summer). Politically unnoticed, in early October in a pronounced outbreak with *R*_*t*_ = 2.5 and an average incidence up to 16,000 cases per day started. After this wave already shows significant decay at the end of October, a lock-down light is imposed. This is reinforced with very restrictive measures at the end of November, although the event had already decayed very strongly. Consideration of incidence alone may have suggested this. Strong reactions that briefly put a lot of strain on the health system followed the anouncement of the December-Lockdown at the end of November 2020. The small spike as a result of New Year’s Day could have been to be expected.

In addition to the simulated waves we draw hyperbolas where (Incidence *R*_*i*_) is constant. This could be recocgnized as a kind of Spreading Index.

This type of plot does not only look at the incidence alone, but also, in conjunction with the new *R*_*i*_, at the entire pandemic dynamic in a very efficient way.

In Fig.8 some points stand out strikingly, which we want to illuminate more closely and also give suggestions for an interpretation. In any case, however, the times of the outbreaks of several waves are quite clearly recognizable. They cannot be seen when concentrating on the incidence axis alone. In the following graph, some points are labeled, on some of them we want to focus briefly.

**Figure 8:**
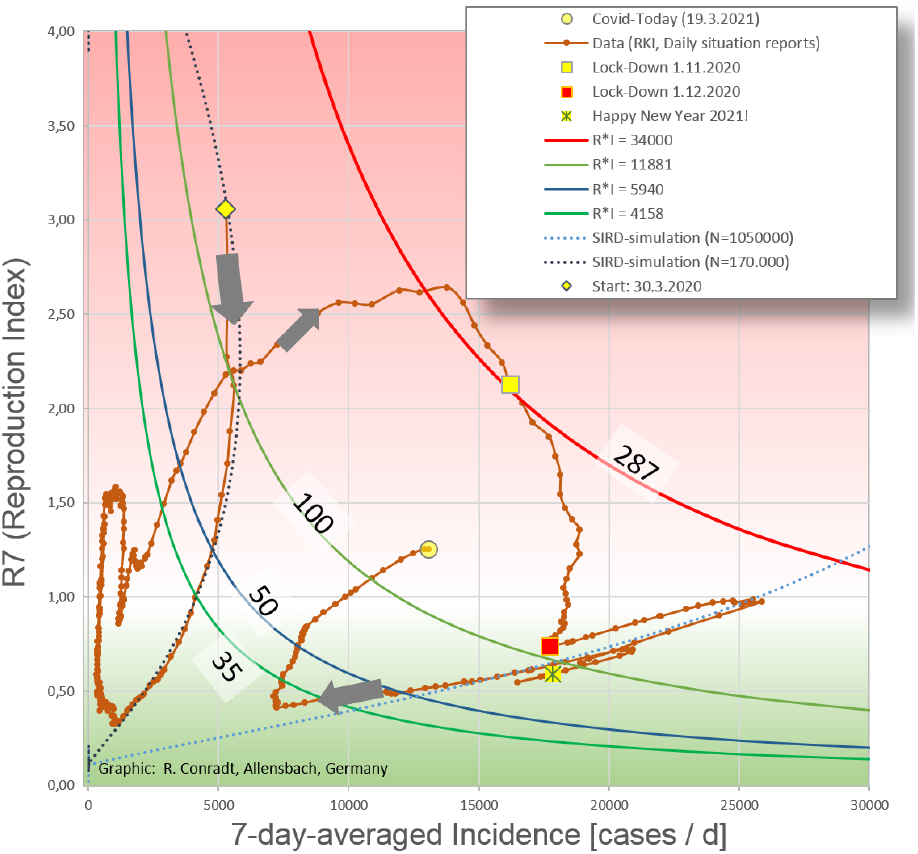
Indicator for Infectious Dynamic. Plotting R_i_ vs. Incidence make distinct features visible. Auxiliary lines, hyperbolas of constant development of the pandemic are drawn. The labels at the hyperbolas mean e.g. “100 cases per 100.000 inhabitants in 7 days at R_i_ = 1”. Corresponding for the 50-and 35-label. In addition, the trajectories of the first wave in 2020 and another unaffected wave are plotted. Distance between the dots is 1 day.

The following paragraphs take up some of the points highlighted in Fig.9 in chronological order. Prior to this, the first wave of infections in 2020 (see Fig.3) passed through. This part of the trajectory follows very closely the simulation and is not further discussed here.

**Figure 9:**
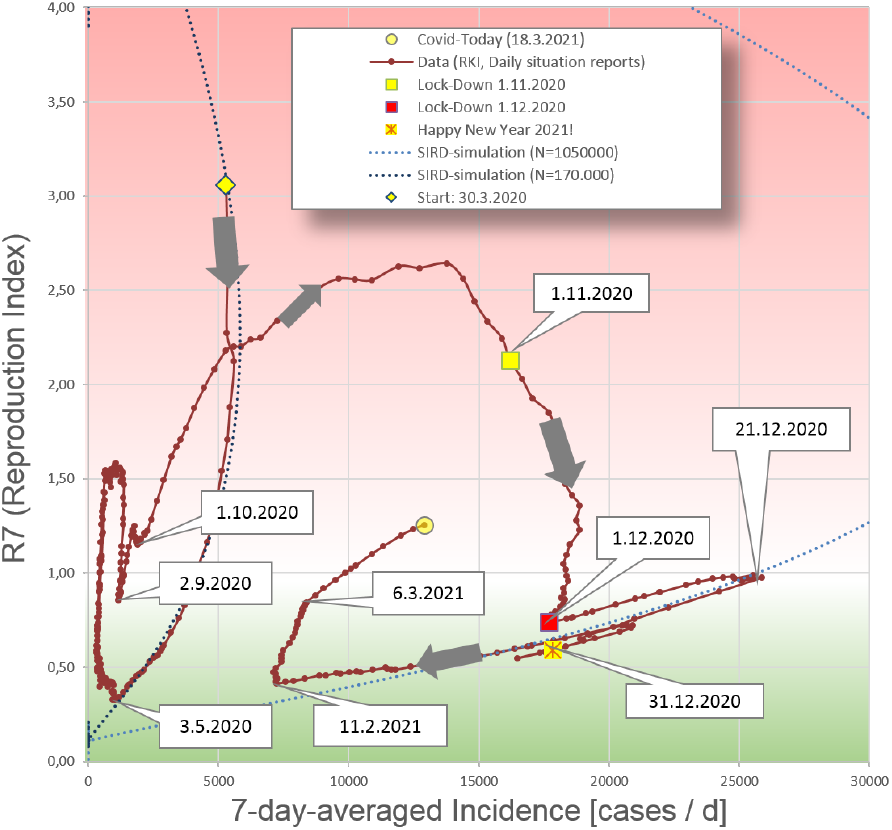
*R_i_* vs. Incidence, obvious features marked. Individual distinctive signatures are marked with their corresponding date. The markings are taken up again in the text as a reference.

**3.5.2020 to 1.10.2020** For May 6 2020, the German Chancellor and the Conference of Minister Presidents decided to relax public life. This concerned opening of stores, restaurants, cultural institutions and museums.

Because of little change in incidence in both, spring and end of summer, it was not noticed that the Reproduction rate was undergoing strong movements. However, these strong changes were only indicated by the Reproduction index *R*_*i*_, but not by RKI’s Reproduction number *R*_*t*_.

**1.11.2020 to 31.12.2020** After the incidence rose very sharply in the fall of 2020, the German government decided on a lockdown light from November 2. This was to be associated with the hope of easing at Christmas.

Since the decrease of the Reproduction rate is not reflected by *R*_*t*_, it was not noticed that the situation has relaxed since the end of October with respect to the Reproduction index *R*_*i*_. Possibly the population, warned by the daily news, reacted itself. Since policy decisions were based mainly on incidence values, a lockdown light was first put into effect from Nov. 2nd. On Dec. 1st a hard lockdown was announced starting Dec. 16th. This hard lockdown provided for widespread closures of businesses, schools, etc. In particular, the relaxations for Christmas that had been promised were withdrawn. This announcement led to a short-lived but sharp increase in infections as many people were squeezed through the eye of a needle in both time and space for their Christmas shopping. The situation eased with the simultaneous start of the Christmas vacations throughout Germany on Dec 22nd 2020.

The small rash on New Year’s Eve was to be expected.

The special thing about the winter until 1.12.2020 was that the felt situation did not correspond to the situation announced in the daily news but rather to the relaxation seen in Fig.11. As a result, the acceptance in the population towards political measures decreased strongly. This can be a danger at the moment when strong restrictions are actually necessary - which may not be accepted by the population.

**Figure 10:**
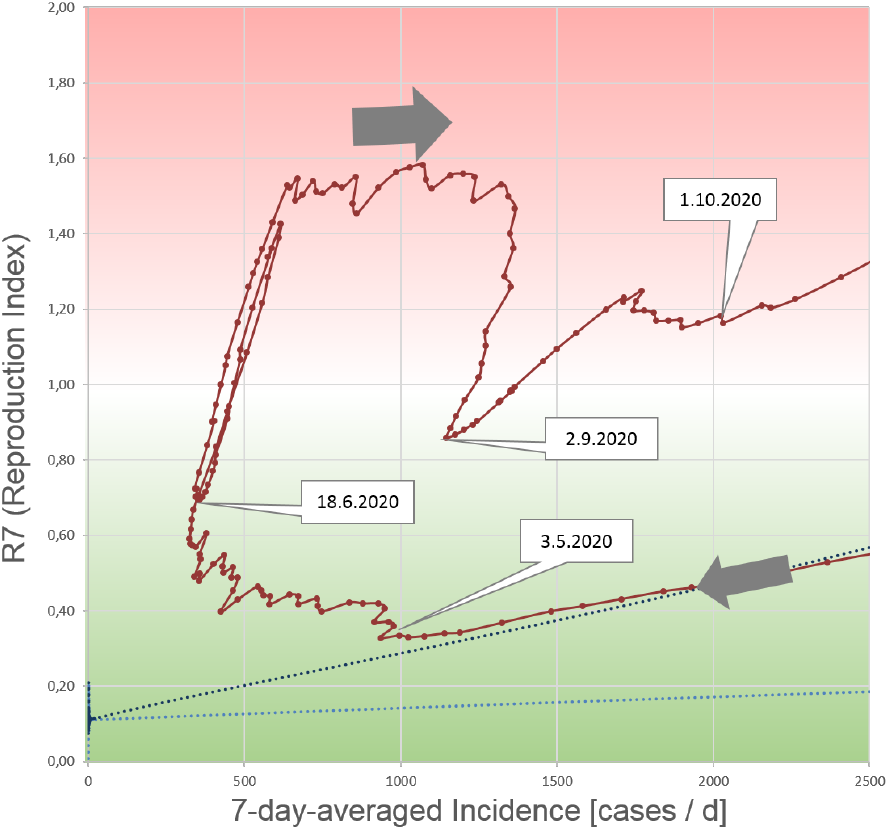
Spring and summer 2020. After 3.5.2020, loosenings came into effect. 6.18.2020: Covid19 outbreak at the Tönnies/Gütersloh company. Summer vacations end in early to mid-September.

**Figure 11:**
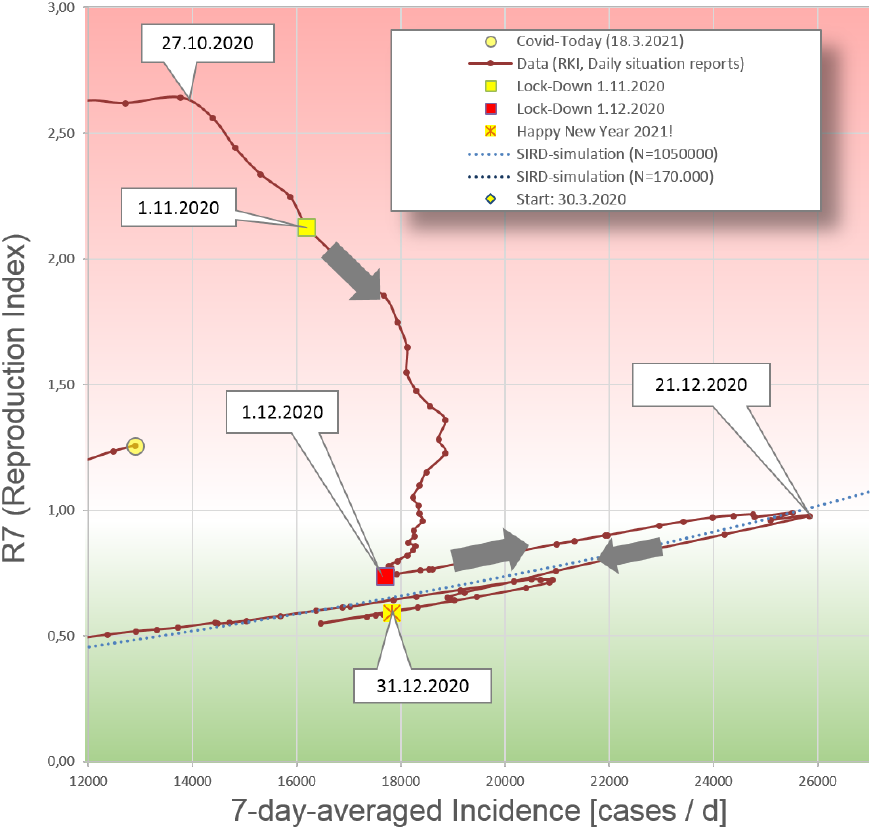
Fall and winter 2020. While the Reproduction index R_i_ was already falling considerably, a lockdown light was put into effect on 2.11.2020. A hard Lockdown was announced on 1.12. for the time starting from 16.12. German Christmas vacations start on 22.12. The small rash on New Year’s Eve was to be expected.

**11.2.2021 up to now** Under the influence of the still ongoing lockdown from 16.12.2020, both incidence and Reproduction index *R*_*i*_ continued to decrease. From 11.2.2021, incidence values stagnated and did not drop further. A simultaneous sharp increase in the Reproduction index *R*_*i*_ was not noticed, as the RKI Reproduction number *R*_*t*_ continued to hover around the value of “1” and showed only a small abnormality. By the way: the same kind of peak-event as in Streeck’s study[11].

**Figure 12:**
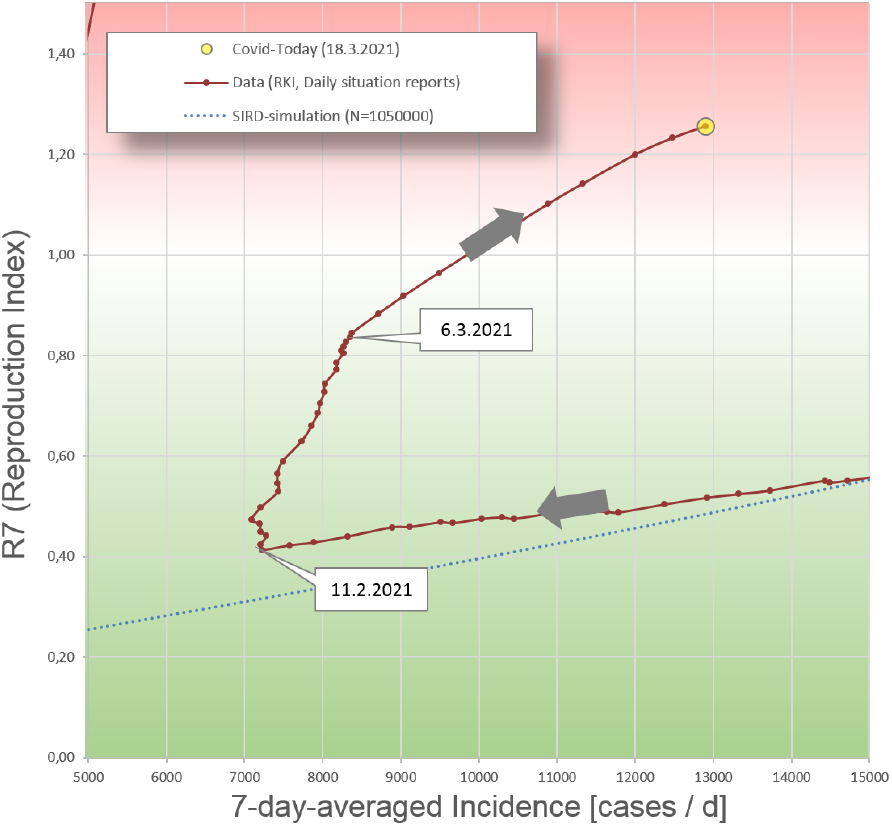
Start of 3rd wave. With the 11.2.2021 (“Weiberfasnacht” in Germany) the decrease of the incidence values stagnates and at the same time the Reproduction index R_i_ increases.

For completeness, the same kind of diagram is shown again, but this time RKI’s *R*_*t*_-values are used instead:

Finally we can state that indroducing a new Reproduction index *R*_*i*_ can provide a lot of additional information in with respect to the RKI’s *R*_*t*_.

The method of projecting the pandemic data onto two different layers yields a much more meaningful insight into pandemic events than projection onto the incidence axis alone. This proposed way of plotting only shows information when using the new proposed Reproduction Index *R*_*i*_.

The way of using *R*_*i*_ and displaying pandemic data is not limited to the present Covid19-pandemic. We assume this to be a powerfull tool for monitoring future pandemic to keep them under immediate control.

It would be very helpfull and easy to generate a map with spatially resolved indication of a kind of “Spreading Index” which is given by *R*_*i*_ *CaseNumbers*. In such map pandemic growth is immediately visible.

**Figure 13:**
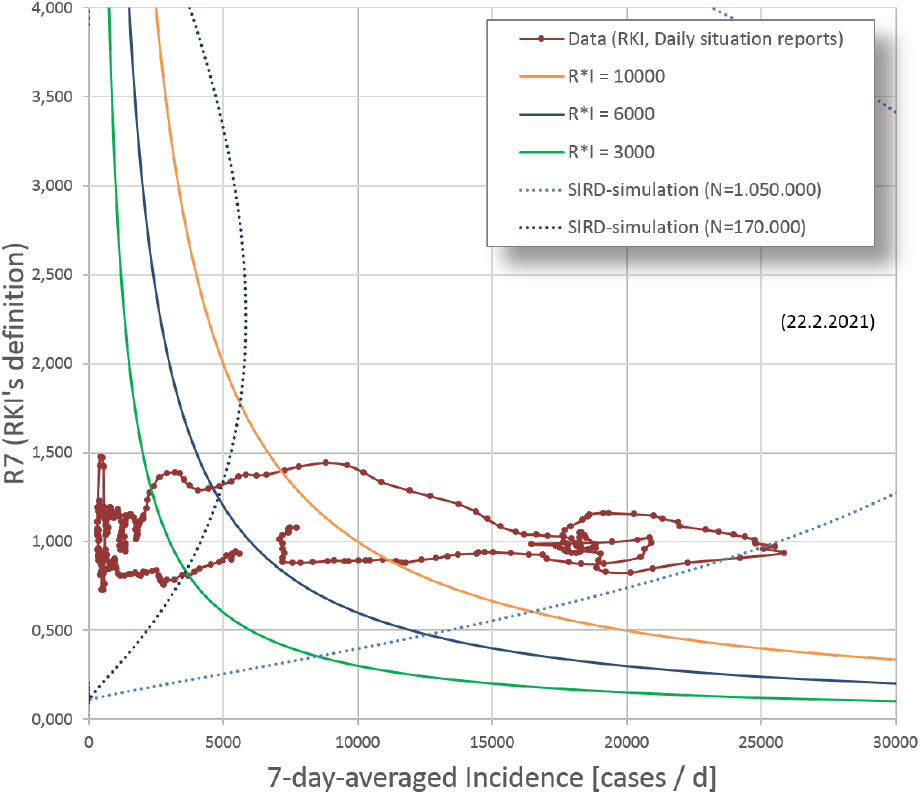
*R_t_* vs. Incidence. does not show any typical signature compared to Fig.8 when the value according to Eq.(13) is used for R. De facto, it corresponds to a projection of Fig.8 onto the Incicence axis

The pronounced signatures in the (*R*_*i*_ | *I*)-plots give the possibility to use a Lock-Down for a short interrupt - find the infected individuals and continue the life with minor restrictions. This makes a lockdown a short-term tool to find infected individuals. This avoids using a hard lockdown as a permanent measure to dry up a pandemic.

This paper is dedicated to all those who died in the pandemic, especially to those who passed away in loneliness.

Allensbach, 28.3.2021, Robert Conradt

## Many thanks to…

… Prof. Paul Leiderer for the great support, a lot of discussions and the multiple review of the manuscript. Dear Paul - thank you very very much!!! I would like to take this opportunity to thank Prof. Stephan Herminghaus for the encouraging and spontaneous midnight discussion. Thank you, Stephan!

Getting the ball rolling in March 2020 I, was supported by my classmate Prof. Christoph Klein. Thank you very much, Christoph!

My good friend Dr. Armin Lambacher was a very great help in reviewing the first manuscripts. Dear Armin - thank you very much!

## Data Availability

All data were published by Robert-Koch-Institute

https://www.rki.de/DE/Content/InfAZ/N/Neuartiges_Coronavirus/Situationsberichte/Gesamt.html

## 3 Supporting Information

### 3.1 The Reproduction Index *R*_*∞*_

Using the set of equations Eq.(4) we combine *R* and *D* to a new *R*^*′*^. This is the population which has left the population *I* with the rate *m*_0_ + *r*_0_. For the duration of incectiosity Δ*t* we use (*m*_0_ + *r*_0_)^*−*1^. We result in a *SIR*^*I*^-model with a graph for the Reproduction:

**Figure 14:**
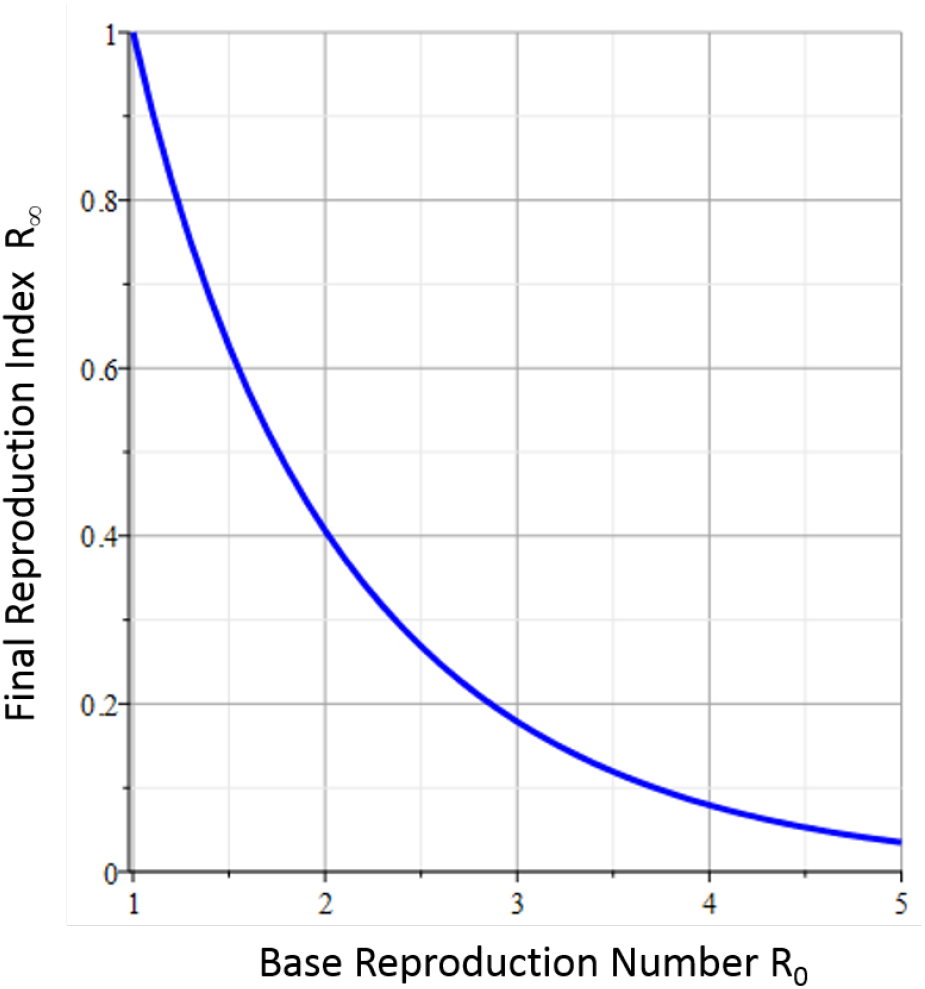
Final Reproduction Index *R*_∞_. only depends on the base Reproduction index R_0_.

At the end of a pandemic the Reproduction Index decays to a number which is given by the following two expressins which can be easily derived from the DES Eq.(4):

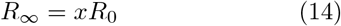

together with a transcendental equation for *x*:

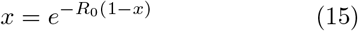

### 3.2 Determination of the parameters for the SIRD model

From the data of the daily situation report of the RKI [9] the parameters of the SIRD model (see Fig.1) can be obtained^18^.

#### 3.2.1 Infection rate *c*

**Figure 15:**
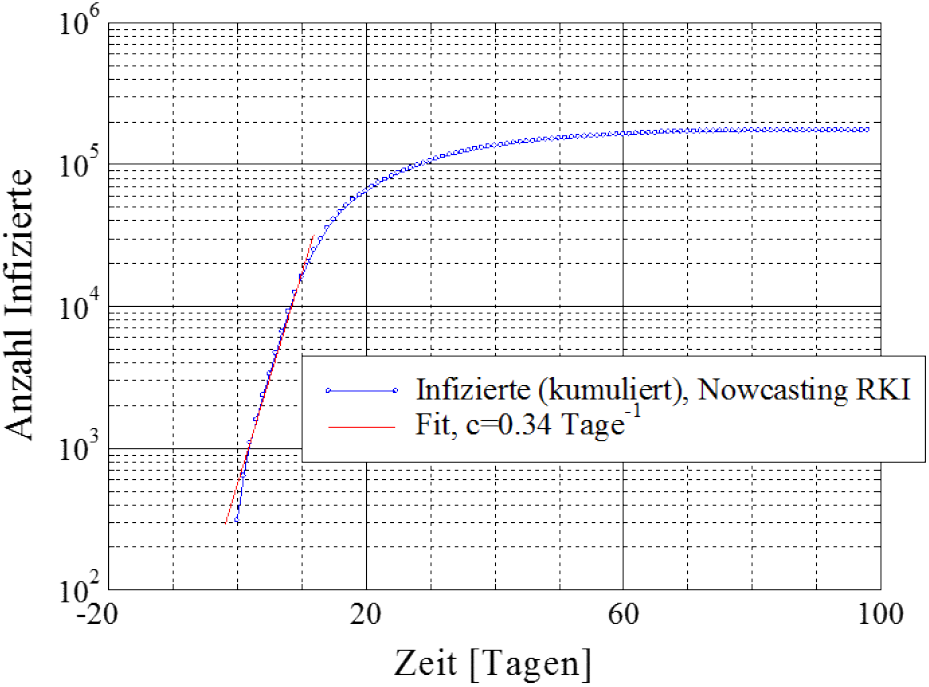
To get the infection rate c: From the exponential onset the infection rate can be determined. With a duration of infectivity of Δt=12 days, a basic Reproduction index results in R_0_=4.0

#### 3.2.2 Recovery rate *r*_0_

**Figure 16:**
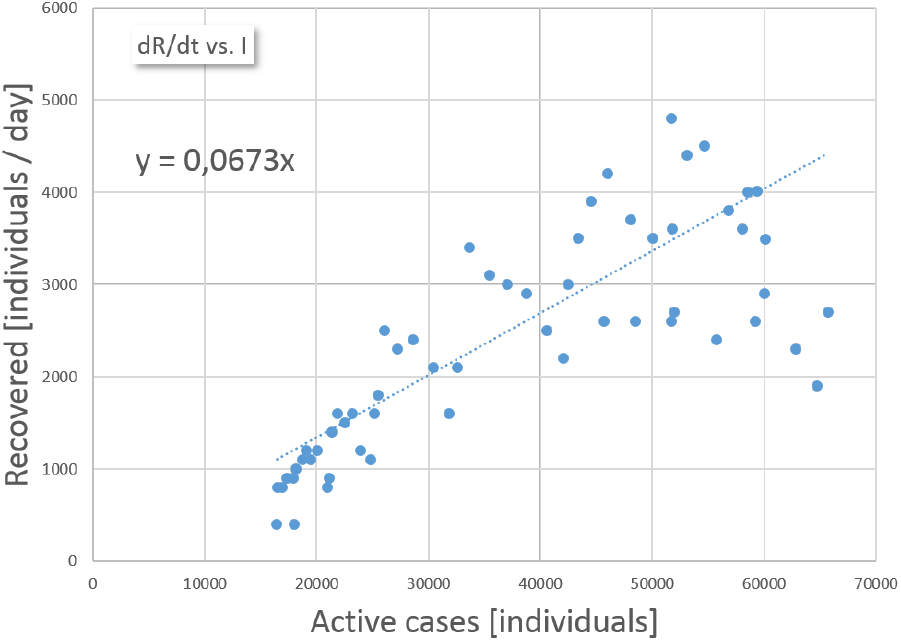
Determination of the recovery rate *r*_0_: The values of the increase in recovered persons are plotted against the infected persons existing at the same time. A value of r_0_=0.067 recovered persons per day and infected person is determined. Note that the number of recovered individuals is an estimation by RKI.

#### 3.2.3 Mortality rate *m*_0_

**Figure 17:**
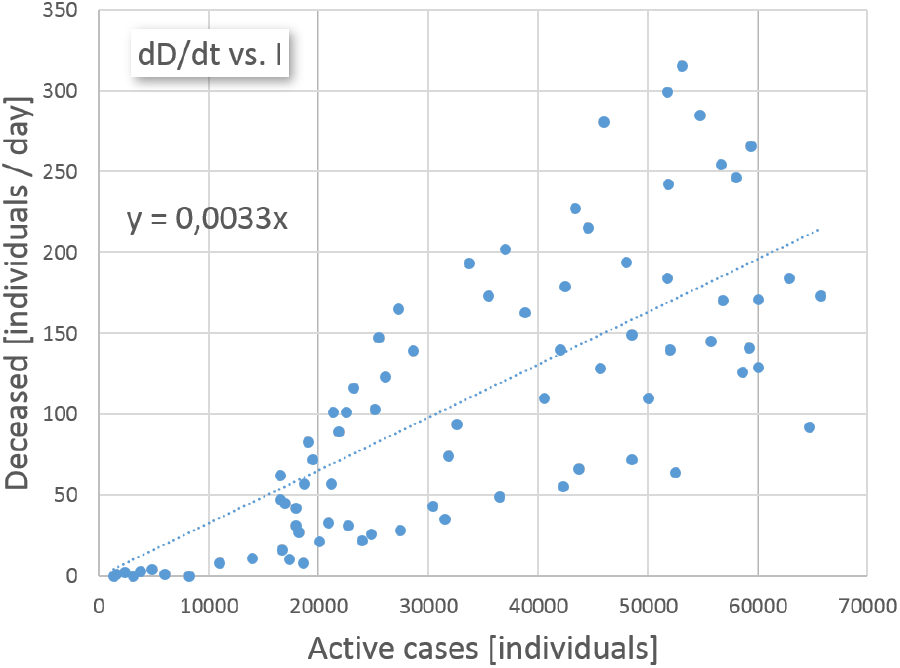
Obtaining the mortality rate *m*_0_: The values of daily deceased persons is plotted against the infected at the same time. A mortality rate of m_0_=0.0033 day^−1^ is found.

#### 3.2.4 Nowcasting vs. daily situation reports

The RKI releases the Reproduction number with the definition according to Eq.(13) based on the data of the so-called nowcasting [8]. The new infections are not assigned to the date of their reporting - as in the daily situation reports - but to the reconstructed date of the actual infection. This makes these data more reliable and valuable.

**Figure 18:**
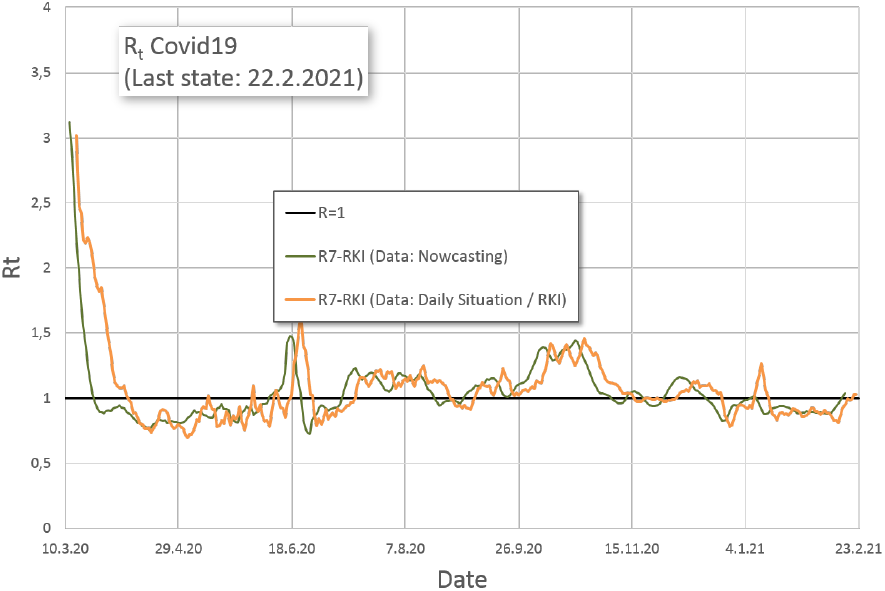
Comparison *R*_t_: Nowcasting and Daily Situation Report. Both curves for R_t_ of the RKI are performed according to the same definition Eq.(13). The nowcasting data are subsequently checked and corrected if necessary. As a result, this curve appear be partially shifted to earlier dates to be smoother

## Footnotes

2 For nomenclature: *R*_*i*_ (Reproduction index) denotes the new index, *R*_*t*_ (Reproduction number) denotes the definition of the RKI. The general term “Reproduction Rate” refers to both numbers (*R*_*i*_ and/or *R*_*t*_).

3 Herd immunity is achieved through the pandemic itself or by vaccination.

4 distance, viral load, contact-duration, the immune response of the infected person, etc.

5 Since around October 2020, a value for *I* will be specified explicitely. Before, *I* could be derived from the totaled new infections minus the recovered *R* and deceased *D* individuals.

6 values for mortality *m*_0_ and recovery rate *r*_0_ can be obtained independently, values for *c* and *N*_*ges*_ are obtained by plotting the rate according to Eq.(1) against the number of actively infected *I*,

7 cases (new or total), deceased persons (new or total), recovered persons (new or total). Self-consistency of the data can be checked by summing up the new counts and comparing them to the totals.

8 This is in the middle of the mentioned 12.3 days (see Fig.2) and the 14.2 days derived from (*r*_0_ + *m*_0_)^*−*1^

9 In the 7-day R value, both sums are shifted by 4 days, but extend over 7 days each for even better smoothing.

10 corresponding to the maximum of Active Cases in Fig.3

11 Also here the tiny shift between the two curves arise from principal time-shift between *R*_*t*_ and *R*_*i*_ and additionally from the subsequent assignment of some cases to the correct infection date in the RKI’s Nowcastig-data

12 Note that RKI’s *R*_*t*_ does not reproduce this observation.

13 One easily can derive from the differential equations that the proposed coordinates can be derived from the coordinates SI(R+D) directly.

14 marked as “Covid19 (RKI, Daily Status)” in the legend.

15 This is not completely correct, because for the calculation of *R*_*i*_(*t*) there is a time offset (generation time) between incidence and prevalence

16 here for 35 new infections in 7 days per 100,000 inhabitants, for 50 and for 200. The values are based on the population of Germany with currently 83,166,711 inhabitants

17 Assuming that all intensive care beds can actually be used

18 as of June 2020

## Notes

### Competing Interest Statement

The authors have declared no competing interest.

### Funding Statement

The work was financed exclusively by own means

